# One year later: Tracking the continued growth of mental illness stigma in England

**DOI:** 10.1101/2025.10.10.25337746

**Authors:** A. Ronaldson, C. Henderson

## Abstract

Using data from the Attitudes to Mental Illness (AMI) survey, we previously reported positive change in mental health stigma in England between 2008-2019. However, following the conclusion of the Time to Change campaign in 2021, 2023 data revealed a deterioration in several stigma-related attitudes. This report presents AMI survey 2024 results, examining changes over the past year. Regression analyses assessed stigma-related knowledge (Mental Health Knowledge Schedule (MAKS)), attitudes (Community Attitudes toward the Mentally Ill scale (CAMI)) and behavioural intent (Reported and Intended Behaviour Scale (RIBS-IB)), along with willingness to interact based on vignettes of depression and schizophrenia. The proportion of respondents achieving 2023-level MAKS and CAMI scores declined significantly (by 3.5%, p=0.028; and 7.0%, p<0.001), while RIBS-IB scores showed a non-significant decrease. Vignette responses remained stable, but there are signs of increasing desire for social distance. This report explores potential drivers of these trends.

## Background

Public stigma towards people with mental illness comprises the beliefs, attitudes and behaviour towards people perceived as having, or with a diagnosis of, a mental illness. We previously found evidence that public stigma in England worsened over: 1994-2003, especially 2000-2003(1) as measured by general population surveys of attitudes to people with mental illness in general; and from 2019-2023(2), as measured by stigma-related knowledge, attitudes and intended behaviour pertaining to people with mental illness in general. There was a significant improvement in these three outcomes between these periods, particularly between 2008-2019(3). The Time to Change stigma reduction programme(4, 5) ran 2009-2021, including a social marketing campaign aimed at adults aged 25-44 in middle to lower middle-income groups, and work with several target groups, and is likely to have contributed to this improvement(6).

Other aspects of stigma measured during this period are available for comparison. For example, consistent with general population surveys, surveys of discrimination experienced by people using mental health services undertaken between 2008-14 showed a significant reduction overall and in numerous life areas: friends, family, dating, finding a job, keeping a job, police, education, religious activities, social life in general, privacy, and starting a family (7). General population surveys measuring desire for social distance from people described in vignettes with symptoms of depression and schizophrenia also showed improvements between 2007-2021(2). This improvement was maintained between 2021-2023(2), in contrast to the deterioration in public stigma measured with scales comprising items about people with mental illness in general.

This divergent pattern of change suggests that behavioural, interpersonal intentions towards known individuals should be distinguished from attitudes and beliefs towards people with mental illness in general, rather than equating public stigma with all interpersonal stigma(8). To continue tracking general population stigma, we therefore aimed to compare each of these aspects of stigma between 2023-2024.

## Methods

The Attitude to Mental Illness (AMI) survey has been carried out in England since 2008, with the latest in 2024. A quota sampling frame was used to ensure a nationally representative sample of adults (16 years or older) living in England, respondents were not resampled in later surveys, and approximately 1700 respondents took part each time. The survey ran 16th September-10^th^ November 2024. Detailed information about the data source, the sampling methods, and the study measures are published elsewhere(2). The King’s College London Psychiatry, Nursing and Midwifery Research Ethics Subcommittee exempted analysis of these survey data as secondary analysis of anonymised data.

Measures of stigma-related knowledge (Mental Health Knowledge Schedule (MAKS(9)), attitudes to mental illness (Community Attitudes toward the Mentally Ill (CAMI(10)), and desire for social distance (Reported and Intended Behaviour Scale (RIBS(11)) were included. These measures have been included in the AMI since 2008 (CAMI) and 2009 (MAKS, RIBS). For the second time in the AMI, we included vignettes from the British Social Attitudes surveys 2007 and 2015(12) of a common mental health problem (‘Stephen’: depression) and a less common problem (‘Andy’: schizophrenia). Participants were asked social distance questions assessing their willingness to engage with the vignette character across six domains: living next door, socialising, forming a friendship, working together as colleagues, accepting them as a family member by marriage, and trusting them to provide childcare for a relative.

We used multiple regression models to evaluate patterns of change in MAKS, CAMI (including CAMI subscales: ‘Prejudice and Exclusion’; Tolerance and Support for Community Care’), and RIBS Intended Behaviour scores. All models used the standardised scores of the measures as dependent variables, meaning the outputs were interpreted in standard deviation units. Logistic regression models were used to assess change in vignette responses (willing versus unwilling) between 2023 and 2024. Models were adjusted for sociodemographic variables: age, gender, ethnicity, socioeconomic position, and government office region^1^.

Analyses were performed with Stata 18.0 (Stata Corp, College Station, Texas, USA).

## Results

Detailed sample characteristics for the sample up to 2023 were provided in a previous publication^1^. The 2024 sample comprised 1563 participants with the distribution of gender and age group remaining stable over time. Changes seen since the introduction of remote data collection in 2019 (i.e., more participants with professional/managerial occupations, more familiarity with mental health problems) were sustained into 2024.

Figure 1 depicts change over time in total CAMI (plus subscales), MAKS, and RIBS Intended Behaviour scores. Previously, we reported decreases in scores on all scales between 2019 and 2023^1^. Between 2023 and 2024, the proportion of respondents reaching 2023-level MAKS scores reduced significantly by 3.5% (p=0.028). The proportion of respondents reaching 2023-level total CAMI scores decreased by 7.0% time (p<0.001). Analysis of the CAMI subscales showed a 5.7% reduction in the proportion of respondents achieving 2023-level scores on the Prejudice and Exclusion subscale (p<0.001) and a 6.9% reduction on the Tolerance and Support for Community Care subscale (p<0.001). Although the proportion of respondents reaching 2023-level RIBS Intended Behaviour scores declined by 1.9%, this change was not statistically significant (p=0.202).

**Figure 1.**
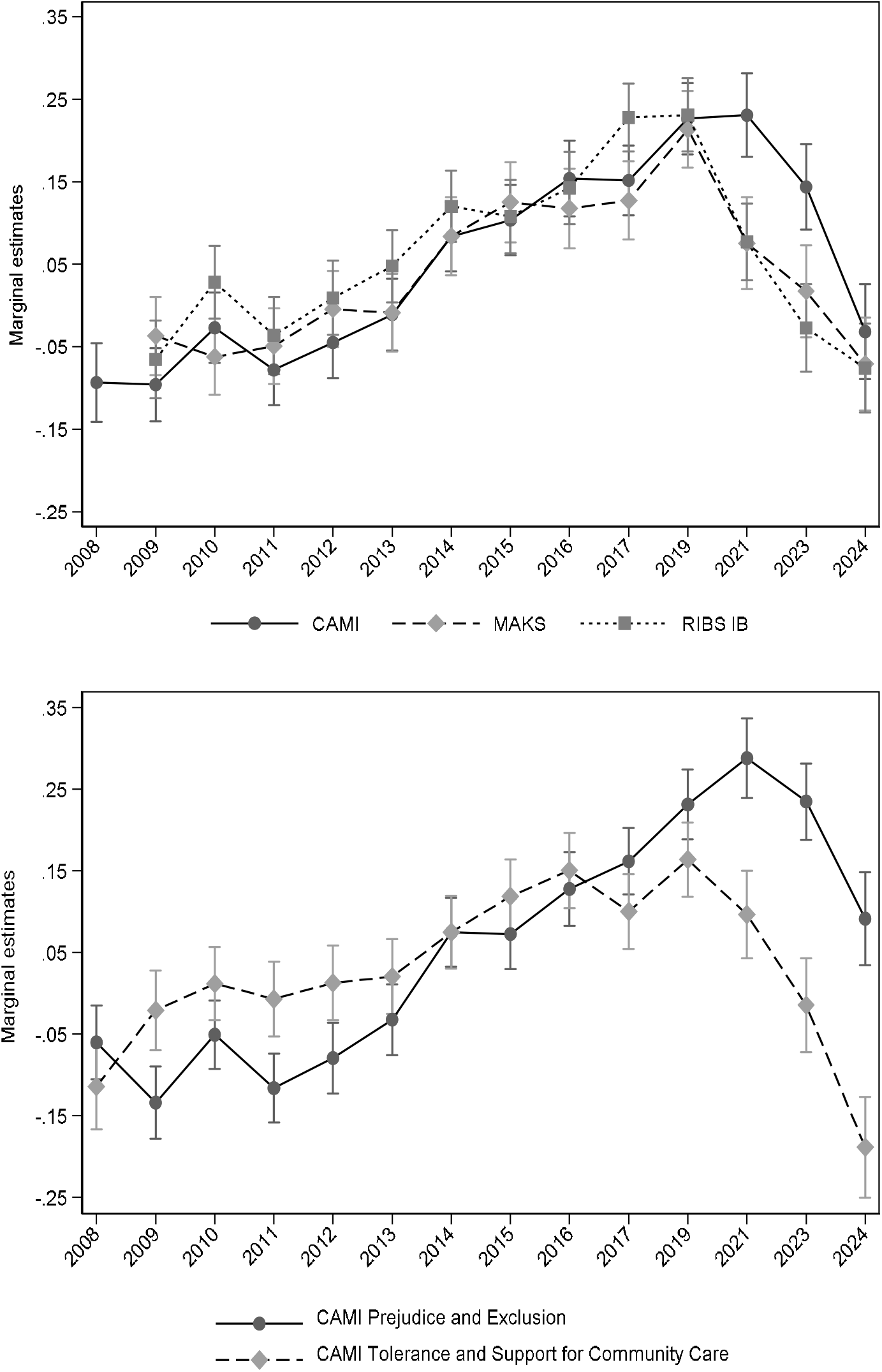
The upper graph presents marginal estimates of stigma-related attitudes (CAMI), knowledge (MAKS), and desire for social distance (RIBS-IB) by year (confidence intervals). The lower graph presents marginal estimates for the subscales of the CAMI scale.

Across vignettes, between 2023 and 2024 the proportion of people who were fairly or very unwilling to interact with ‘Andy’ (schizophrenia vignette) increased across most scenarios with the exception of working together as colleagues and providing childcare. However, fully adjusted logistic regression models showed none of the changes reached statistical significance. Fully adjusted models also revealed no statistically significant changes in unwillingness to interact with ‘Stephen’ (depression vignette), although increases were observed in the proportion of people unwilling to interact across four domains: socialising, forming a friendship, working together as colleagues, accepting them as a family member by marriage, and trusting them to provide childcare for a relative.

## Discussion

Further deterioration in public stigma has occurred over 2023-2024 regarding people with mental illness/mental health problems in general. Desire for social distance from named individuals using vignettes conveying familiarity has not shown significant change over this period, but may increase in future, just as it decreased previously in line with decreases in public stigma(3) and experiences of discrimination(7). There are therefore grounds for concern that people with lived experience of mental ill health will, or may already, experience increasing levels of active discrimination and /or avoidance by others.

Several factors may be contributing to increasing public stigma. Rising levels of mental ill health and help seeking among young people have been interpreted by some public figures as evidence they are ‘work shy’ and misusing the welfare benefits system(13). Economic stressors may lead to greater prejudice towards outgroups. Finally, homicides by people with psychosis which receive intense media coverage and criticism of the mental health services responsible for the care of the perpetrator are likely to influence public stigma(14). A case in Nottingham in 2023 and its coverage has echoes of one in 1992, including the racial stereotyping of Black men(15). However, given delays of several weeks to inpatient admissions in some areas of England, public anxiety about visibly unwell people in their community is increasingly likely and should not be dismissed simply as public stigma. Responsive services are needed to people in crisis and people need to know whom to contact for a response. From a systems perspective(16), greater help seeking and rising levels of mental ill health are disruptive, leading either to greater investment in services or to a backlash in the context of an inability or unwillingness to invest. That the latter may be occurring may reflect economic and labour market circumstances and/or a failure to address structural stigma.

## Acknowledgements

We are grateful for the collaboration on the evaluation by Kerry Goddard and Anita Fernandes at Mind and acknowledge the valuable contributions of George Hoare and Alex Viccars during their previous roles at the organisation.

## Data availability

The data that support the findings of this study are available from the corresponding author, A.R., upon reasonable request.

## Author contributions

A.R. conducted the data analysis and created the figure. A.R. and C.H. drafted and reviewed the manuscript.

